# Citation Impact and Public Attention Analysis of Benign Prostatic Hyperplasia Research in the Urological Literature

**DOI:** 10.64898/2026.09.01.26361260

**Authors:** Georges (Camille) Motchoffo Simo, Anael Rizzo, Kevin B. Muy, Nancy Quintanilla, Kymora B. Scotland

## Abstract

**Objective:** To determine whether the most cited and the most publicly discussed benign prostatic hyperplasia (BPH) literature describe the same body of work, and whether clinicians and patients read different evidence.

**Methods:** Four Boolean Web of Science searches and four matched Altmetric Explorer searches were run in February 2024, restricted to literature indexed with urologic terminology, and screened in duplicate. Two arms were assembled: the 50 most-cited articles (citation census May 2024) and the 50 with the highest Altmetric Attention Scores. Funding source and intervention focus were hand-coded from the full text of all 100 articles; open-access status came from Unpaywall. Arms were compared with Mann-Whitney U and Fisher exact tests

**Results:** Eight of 50 articles (16%) appeared in both arms. Citation-selected articles were older (median 2008 versus 2018, p<0.001), more often randomized trials (58% versus 22%, p<0.001), and more often published in urology-specific journals (96% versus 66%, p<0.001). Industry funded 50% versus 18% of articles (p=0.001) and non-industry sources 8% versus 36% (p=0.001). Only 10% of citation-selected articles were open access versus 56% (p<0.001). Attention data were recoverable for only 13 citation-selected articles (median score 9 versus 17). The two arms were cited in the 2026 AUA BPH Guideline at indistinguishable rates (22% versus 20%, p=1.00).

**Conclusions:** These are largely distinct bodies of work, separated most decisively by whether they can be read without a subscription, yet both inform guideline development equally. Patients arrive with evidence systematically different from, and no less guideline-relevant than, that underpinning their urologist’s training.

## Introduction

Benign prostatic hyperplasia (BPH) is among the most common conditions in adult urology, affecting most men beyond the seventh decade and accounting for a substantial share of ambulatory urologic care.^1^ Contemporary management of BPH has evolved to encompass pharmacologic combination therapy, laser enucleation and vaporization, and an expanding class of minimally invasive device-based interventions.^2–4^

That literature is large, heterogeneous, and unevenly familiar even to specialists, and bibliometric analysis has become a standard method for mapping it. By ranking publications according to citation frequency, it identifies the work that has most shaped subsequent scholarship and yields a defensible reading list for trainees. The method has been applied across urologic subspecialties,^5^ and its assumptions are well understood:^6^ citation counts accrue slowly, reward methodological rigor and multi-institutional scale, and reflect the judgment of researchers rather than of patients or the public.^7,8^ It has recently been applied to BPH itself, in an analysis of 2,613 publications on surgical management indexed between 2016 and 2025.^9^

Altmetric was developed to capture a different signal.^10^ Altmetric indicators aggregate mentions in news outlets, policy documents, reference managers, blogs, and social media platforms, producing a weighted Attention Score intended to reflect the reach of a publication beyond the academy.^11,12^ Whether that signal is a complement to citation impact or a substantially different construct remains contested.^13,14^

This distinction has acquired clinical weight. Patients with lower urinary tract symptoms (LUTS) increasingly arrive having researched their condition and request specific interventions by name: prostatic urethral lift, water vapor thermal therapy, prostatic artery embolization, aquablation, or plant-derived supplements such as saw palmetto. The urologist’s ability to respond depends on familiarity with the evidence the patient has encountered. If citation-selected and attention-selected literatures overlap substantially, that familiarity can be assumed. If they do not, clinician and patient may be reasoning from different evidence bases without either recognizing it.

We therefore conducted a paired bibliometric and Altmetric analysis of the BPH literature with three aims: (1) to characterize the most cited and most publicly discussed publications on BPH management; (2) to determine how far these two bodies of work overlap and how they differ; and (3) to assess whether either is preferentially reflected in contemporary clinical practice guidelines.

## Methods

### Study design

This cross-sectional bibliometric study employed a dual-database design: the most-cited BPH articles indexed in the WoS, and the articles with the highest Altmetric Attention Scores in Altmetric Explorer. Because the study analyzed only publicly available bibliographic data and published articles, institutional review board approval was not required.

### Search strategy

The search strategy was developed with research librarians at an academic institution and informed by expert input from a board-certified endourologist (K.B.S.). Five term categories were selected: general condition, surgical management, medical management, advanced equipment, and urologic terms.

The terms were combined with the field tag TS using OR within categories and AND between them, rendering four queries: surgical management, medical management, advanced equipment, and combined surgical and advanced equipment. Every query required a urologic term, a deliberate restriction whose consequences are addressed in the **Limitations**. Full term lists and query syntax are in **Supplementary Methods**. Both databases were searched in February 2024 with Altmetric queries converted to PubMed-equivalent syntax. The 100 highest-ranked records per WoS query were exported; the four Altmetric queries returned 367 records and 252 unique outputs after deduplication. The citation census date was 22 May 2024.

### Screening and selection

Records were uploaded to Rayyan for duplicate removal and blinded screening (**Figure 1**). Three reviewers (A.R., K.M., N.Q.) independently screened titles and abstracts; an article was retained when at least two reviewers approved it, with discrepancies resolved by consensus and adjudication by the senior author. The citation arm was restricted to primary research articles in English. The attention arm applied deliberately broader publication-type criteria, permitting systematic reviews, meta-analyses, and narrative reviews, because these formats generate a disproportionate share of online attention and excluding them would have misrepresented the construct under study. All other exclusions, including animal, in vitro, pediatric, and prostate malignancy studies, were applied identically to both arms.

### Data extraction

Extracted variables comprised publication year, journal, journal impact factor at publication (Journal Citation Reports),^15^ citation count, author count, country of first-author affiliation, study design, institutional scale, and funding source.

Funding source was determined by reading the full text of all 100 articles and was coded from each article’s funding or financial-support statement into four mutually exclusive categories: industry (commercial sponsorship, sponsor-funded medical writing, or authorship by an employee of a commercial entity), non-industry (governmental, institutional, professional society, or foundation support), not funded (an explicit statement that the work received no funding), and not reported (absence of any funding statement). Author conflict-of-interest disclosures were not treated as funding statements. Institutional scale was determined from the author affiliations of every article, including reviews and guidelines, and coded as single institution where all authors shared one institution.

Open-access status was determined by DOI lookup in Unpaywall in August 2026 for the citation arm and from Altmetric Explorer for the attention arm.^16^ Attention sources were grouped by audience as public-facing (news outlets, Wikipedia, Facebook, video, Reddit), professional-leaning (Twitter/X, blogs), and policy documents. Thirteen of the 50 citation-selected articles were retrievable in the Altmetric export; because non-retrieval reflected query matching rather than absence of online attention, no values were imputed and attention was not used as an outcome measure in the citation arm.

Intervention focus was assigned to one of six mutually exclusive categories by keyword derivation followed by manual verification of all 100 articles (**Supplementary Methods**). Each article was cross-referenced against the reference list of the 2026 American Urological Association guideline on the management of LUTS attributed to BPH.^2–4^

### Statistical analysis

Continuous variables were summarized as medians with interquartile ranges (IQR) and compared using the Mann-Whitney U test, reflecting the non-normal distribution of citation data. Categorical variables were summarized as counts with percentages and compared using Fisher exact tests. Associations between continuous variables were assessed with Spearman rank correlation. Statistical significance was set at α = 0.05. Analyses were performed in R version 4.6.1.

## Results

### The most-cited BPH literature

The 50 most-cited articles accumulated 8,621 citations (median 152, IQR 106 to 211; range 79 to 563) and were published between 1993 and 2018, with a median publication year of 2008 (IQR 2003 to 2012). Publication was concentrated: European Urology accounted for 29 articles (58%) and Urology for 16 (32%), and 48 (96%) appeared in urology-specific journals.

Randomized controlled trials predominated (29, 58%) and 31 articles (62%) were multi-institutional. Journal impact factor did not predict citation accumulation within this already-influential set (Spearman rho = 0.098, p = 0.50), although publication year and impact factor were strongly associated (rho = 0.738, p < 0.001). Full descriptive data appear in **Supplementary Table S1**.

### The most publicly discussed BPH literature

The 50 attention-selected articles had a median Attention Score of 17 (IQR 11 to 36, range 6 to 693) and a median publication year of 2018 (IQR 2012 to 2021). Retrospective and observational studies were most common (20, 40%), followed by randomized trials (11, 22%) and systematic reviews or meta-analyses (10, 20%). Thirty-three articles (66%) appeared in urology-specific journals.

### What the attention signal measures

Because the Attention Score aggregates heterogeneous sources, we decomposed it to determine which audience it reflects. News outlets contributed 71.5% of the total score across the attention arm, Twitter/X 20.0%, blogs 3.7%, Wikipedia 2.6%, and policy documents 1.9%. Grouped by audience, public-facing sources accounted for 74.4%, professional-leaning sources for 23.7%, and policy documents for 1.9%. The arm attracted 338 news mentions across 25 articles and 24 policy citations across 10 (**Figure 2**). A professional signal was nonetheless present: all 50 articles had Mendeley readers, totaling 4,141, a channel that does not contribute to the Attention Score. The two signals coexist but are measured separately, and we therefore interpret the attention arm as the BPH literature circulating in the public sphere rather than as a measure of clinician readership.

### The two literatures compared

Eight articles (16%) appeared in both arms, and 13 of the 50 most-cited articles (26%) had any recorded presence in the Altmetric export. Beyond that overlap the arms diverged on nearly every measured characteristic (**Table 1**).

**Table 1.** Characteristics of the citation-selected and attention-selected BPH literature.

| Characteristic | Citation arm (n=50) | Attention arm (n=50) | P value |
| --- | --- | --- | --- |
| **Access** |  |  |  |
| Open access, n (%) | 5 (10) | 28 (56) | **<0.001** |
| **Publication characteristics** |  |  |  |
| Year of publication, median [IQR] | 2008 [2003–2012] | 2018 [2012–2021] | **<0.001** |
| Journal impact factor, median [IQR] | 6.5 [2.2–10.5] | 4.8 [2.6–7.4] | 0.42 |
| Number of authors, median [IQR] | 7 [6–9] | 7.5 [5–10] | 0.95 |
| Urology-specific journal, n (%) | 48 (96) | 33 (66) | **<0.001** |
| **Study design** |  |  |  |
| Randomized controlled trial, n (%) | 29 (58) | 11 (22) | **<0.001** |
| Prospective, n (%) | 5 (10) | 3 (6) | 0.71 |
| Retrospective/observational, n (%) | 16 (32) | 20 (40) | 0.53 |
| Systematic review/meta-analysis, n (%) | 0 (0) | 10 (20) | **0.001** |
| Review/expert opinion, n (%) | 0 (0) | 6 (12) | **0.027** |
| **Study setting** |  |  |  |
| Multi-institutional, n (%) | 31 (62) | 33 (66) | 0.84 |
| Single-institution, n (%) | 19 (38) | 17 (34) | 0.84 |
| **Funding source** |  |  |  |
| Industry, n (%) | 25 (50) | 9 (18) | **0.001** |
| Non-industry, n (%) | 4 (8) | 18 (36) | **0.001** |
| Not funded, n (%) | 6 (12) | 11 (22) | 0.29 |
| Not reported, n (%) | 15 (30) | 12 (24) | 0.65 |
| **Intervention focus** |  |  |  |
| Pharmacotherapy, n (%) | 18 (36) | 9 (18) | 0.070 |
| Laser enucleation/vaporization, n (%) | 18 (36) | 9 (18) | 0.070 |
| Conventional resection, n (%) | 5 (10) | 3 (6) | 0.71 |
| Direct-to-consumer MIST, n (%) | 4 (8) | 8 (16) | 0.36 |
| Phytotherapy/supplements, n (%) | 0 (0) | 5 (10) | 0.056 |
| Outcomes, diagnostics, guidelines, n (%) | 5 (10) | 16 (32) | **0.013** |
| **Guideline uptake** |  |  |  |
| Cited in AUA 2026 Guideline, n (%) | 11 (22) | 10 (20) | 1.00 |

Continuous variables compared with the Mann-Whitney U test; categorical variables with the Fisher exact test. MIST, minimally invasive surgical therapy. Funding was determined from the full text of all 100 articles and coded as industry (commercial sponsorship, sponsor-funded medical writing, or authorship by an employee of a commercial entity), non-industry (governmental, institutional, professional society, or foundation), not funded (an explicit statement that no funding was received), or not reported (no funding statement present). Conflict-of-interest disclosures were not treated as funding. IQR, interquartile range; AUA, American Urological Association.

The largest difference concerned access. Five of 50 citation-selected articles (10%) were open access against 28 of 50 attention-selected articles (56%; p < 0.001), and 42 of the 45 closed-access citation-arm articles appeared in European Urology or Urology. The five open-access citation-arm articles were all studies of minimally invasive or patient-facing topics.

Industry funded 25 citation-selected articles (50%) against 9 attention-selected articles (18%; p = 0.001), whereas non-industry sources funded 4 (8%) against 18 (36%; p = 0.001). Six citation-arm and 11 attention-arm articles were not funded (12% versus 22%, p = 0.29), and 15 versus 12 carried no funding statement (30% versus 24%, p = 0.65). Among articles carrying a funding statement, industry sponsorship accounted for 25 of 35 (71%) in the citation arm against 9 of 38 (24%) in the attention arm (p < 0.001).

Citation-selected articles were older by a median of ten years (2008 versus 2018, p < 0.001; **Supplementary Figure S1**), more often randomized trials (58% versus 22%, p < 0.001), and more often published in urology-specific journals (96% versus 66%, p < 0.001). Systematic reviews and meta-analyses accounted for 20% of the attention arm and none of the citation arm (p = 0.001). Journal impact factor (6.5 versus 4.8, p = 0.42) and author count (7 versus 7.5, p = 0.95) did not differ (**Figure 3**).

### What each literature is about

The arms addressed different interventions. Laser enucleation or vaporization accounted for 18 citation-selected articles (36%) against 9 attention-selected articles (18%; p = 0.070) and pharmacotherapy for 18 (36%) against 9 (18%; p = 0.07), whereas outcomes, diagnostics and guidelines accounted for 5 (10%) against 16 (32%; p = 0.013) and direct-to-consumer minimally invasive therapies for 4 (8%) against 8 (16%; p = 0.36) (**Figure 3**). Phytotherapy was absent from the citation arm entirely: no study of saw palmetto, Serenoa repens or Pygeum africanum appeared among the fifty most-cited articles, against five in the attention arm (0% versus 10%, p = 0.056), reported as a descriptive contrast rather than a formal finding.

### Public attention to the most-cited literature

Attention data were retrievable for only 13 of the 50 citation-selected articles (**Supplementary Table S2**). Within this subsample the median Attention Score was 9 (range 0 to 86) against 17 in the attention arm, seven of the 13 scored below 10, and citation count and Attention Score were uncorrelated (rho = -0.127, p = 0.68). Because these articles were retrieved by a different search route rather than sampled at random, the comparison is descriptive and no values were imputed for the remaining 37. Retrieval was unrelated to attention, since the 13 spanned the full range of scores and five fell below the attention arm’s minimum. In a bounded sensitivity analysis, assigning every non-retrieved article a score below 6 yields a citation-arm median of 0 to 5 against 17, significant under every assumption tested (all p < 0.001). This establishes only that the comparison would not be overturned and not that the 37 articles attracted little attention.

### Guideline uptake

Eleven citation-selected articles (22%) and ten attention-selected articles (20%) were cited in the 2026 AUA guideline on the management of LUTS attributed to BPH (p = 1.00) (**Figure 3**). The two literatures contributed to guideline development at indistinguishable rates despite sharing only 14% of their membership.

## Discussion

The most cited and the most publicly discussed BPH literature are, to a substantial degree, two different bodies of work. They share eight of 50 articles and diverge on publication era, study design, funding source, subject matter, and most consequentially on whether they can be read without an institutional subscription. Only 10% of the 50 most-cited BPH articles are freely available, against 56% of the most publicly discussed. A patient investigating their own condition cannot access 90% of the evidence base on which their urologist’s training rests.

This is not principally a difference of quality:^17,18^ journal impact factor did not differ between the two arms, and the two literatures were cited in the 2026 AUA guideline at statistically indistinguishable rates. Both, in other words, inform contemporary practice. What separates them is visibility: the citation-selected literature is largely industry-funded randomized trial work published behind paywalls in two specialty journals; the attention-selected literature is largely non-commercial synthesis published openly across a wider range of venues. These are different publication economies, and they produce different reading publics.

Because the attention signal in this literature is predominantly public rather than professional, the clinical consequence is specific. Patients present requesting named interventions, and the evidence they cite is drawn from what they were able to read. That the fifty most-cited BPH articles contain no study of *saw palmetto, Serenoa repens, or Pygeum africanum*, while five such studies appear among the most publicly discussed, illustrates the asymmetry, although at this sample size the difference only approaches significance (p = 0.056) and should be read as descriptive. Phytotherapy occupies essentially no space in the literature that shapes urologic training and meaningful space in the literature patients encounter. A urologist who has read only the former is unprepared for a conversation the latter makes inevitable. We do not argue that research priorities should follow public interest, which would be a poor guide to scientific importance, only that a specialty should remain equipped to answer the questions its patients ask.

A similar pattern holds for device-based therapy. Studies of prostatic artery embolization, prostatic urethral lift, and related minimally invasive interventions appeared in both arms, but within the citation arm they were markedly over-represented among the few open-access articles. The most publicly discussed article in the citation arm, a randomized comparison of prostatic artery embolization with transurethral resection published openly in a general medical journal, attracted an Attention Score of 86 against a subsample median of 9. Where patient-facing interventions are studied and published openly, the public finds them.

Comparisons of citation-based and Altmetric-based rankings have generally emphasized that Altmetric-selected literature is newer and only weakly correlated with citation impact.^13,14,19^ Our findings reproduce both observations but suggest that neither is the most informative difference. Publication year, study design, and commercial sponsorship all separated our two arms, yet access status separated them most sharply, and access is the only one of these that a reader outside an institution experiences directly. Analyses that stop at citation counts and attention scores may understate what Altmetric captures: not merely newer or more popular research, but research that can be read at all. With news coverage contributing almost three-quarters of the attention signal, the Attention Score here largely measures what reached the public, and it is precisely the freely available work that did.

A contemporaneous bibliometric analysis reached partly overlapping conclusions from a different direction. Tozsin and colleagues9 characterized 2,613 publications on the surgical management of BPH indexed in Web of Science between 2016 and 2025 and described a shift away from transurethral resection toward laser enucleation and minimally invasive techniques. That analysis was restricted to surgical management, covered a recent decade rather than the full citation record, and did not examine Altmetric attention, open-access status, or the audience reached. It maps the productivity of a field; the present analysis asks who can read it. The convergence extends to individual articles, since the randomized comparison of prostatic artery embolization with transurethral resection ranks among the most-cited documents in their corpus and is simultaneously the most publicly discussed article in ours.

Three implications follow. First, curated reading lists derived from citation counts alone are incomplete guides to clinical practice, because they systematically omit the literature patients bring to the consultation. Second, for authors of patient-facing research, particularly on minimally invasive and complementary therapies, open publication materially determines whether the work reaches the population it concerns.^20,21^ Third, guideline panels appear already to draw on both literatures in similar measure, which suggests that the divergence we describe is a problem of individual clinician familiarity rather than of institutional evidence synthesis.

Trainees entering residency today may have encountered BPH largely through what circulates publicly, and if they arrive already oriented by the more visible body of work the distance between the two literatures may widen rather than narrow with training. This is a hypothesis rather than a finding, since we did not study trainees, but it suggests that curated reading lists make explicit a literature that is otherwise difficult to encounter.

### Limitations

This analysis has several limitations. Every query required urologic terminology in the title, abstract, or author keywords, which restricted the corpus to literature circulating within urology and excluded landmark trials published elsewhere, the Medical Therapy of Prostatic Symptoms trial being the clearest example; the concentration of the citation arm in two specialty journals follows from the same restriction. Citation-based ranking disadvantages recent work, and Altmetric coverage began only in 2011, leaving older articles undercounted.^22^ The Altmetric searches retrieved only 13 of the 50 citation-arm articles, a difference in recall between search routes rather than a property of the articles; if retrieval favors articles attracting coverage, the divergence we report is understated rather than exaggerated, and we make no claim about the remaining 37. Attention was accordingly not a primary outcome. Open-access status reflects availability in August 2026 rather than at publication, the two arms applied different publication-type criteria by design, and non-English publications were excluded throughout.^23^

## Conclusion

The BPH literature that shapes academic urology and the BPH literature that reaches the public share eight of 50 articles and differ systematically in age, design, funding, subject matter, and access. Only 10% of the most-cited articles are freely readable, against 56% of the most publicly discussed. Both bodies of work inform current guideline recommendations to the same degree, so the difference between them is one of visibility rather than validity. Clinicians counseling patients who arrive with specific treatment requests should recognize that those patients may have consulted a real and guideline-relevant evidence base, and one that conventional citation-based reading lists do not contain.

## Figures Legends

**Figure 1. Study flow.**

Parallel screening and selection for the citation arm (Web of Science) and the attention arm (Altmetric Explorer). Four Boolean searches were run in each database; the Web of Science searches returned 100 records each and the Altmetric Explorer searches returned 367 records in total. Fifty articles were retained in each arm.

**Figure 2. Composition of the Altmetric Attention Score and its relationship to citation impact.**

(A) Source composition of the total Altmetric Attention Score across the attention arm, using the published source weightings. (B) The same data grouped by audience. (C) Absolute counts of news mentions, Mendeley readers, and policy citations. (D) Citations against Attention Score for the 13 citation-arm articles retrievable in the Altmetric export; the y-axis is broken between 20 and 80. n = 50 per arm.

**Figure 3. Where the two literatures diverge.**

(A) Funding source, coded from the full text of all 100 articles. (B) Study design. (C) Intervention focus, hand-coded in duplicate. (D) Open access and citation in the 2026 AUA guideline. * p<0.05, ** p < 0.01, *** p < 0.001 (Fisher exact test); n.s., not significant. n = 50 per arm.

## Data availability

The coded dataset generated for this analysis, comprising all extracted variables for the 100 included articles, is available from the corresponding author on request.

## Declaration of generative AI and AI-assisted technologies in the manuscript preparation

During the preparation of this work the authors used Claude (Anthropic) to assist with statistical computation, figure construction, all reviewed and edited by authors who take full responsibility for the content of the published article.

