## Supplemental Materials for "Citation Impact and Public Attention Analysis of Benign Prostatic Hyperplasia Research in the Urological Literature"

**Supplementary Methods**

Search term categories

The search strategy was developed with research librarians at the University of California, Los Angeles and informed by expert clinical input from a board-certified endourologist (K.B.S.). Five conceptually distinct term categories were constructed. Terms were combined with the Web of Science field tag TS (title, abstract, author keywords), using the Boolean operator OR within each category. The verbatim query string for each category is reproduced below.

| **Category** | **Web of Science query string** |
| --- | --- |
| Category 1: General condition | ((TS=(Benign prostatic hyperplasia)) OR TS=(Benign prostate hyperplasia)) OR TS=(BPH) |
| Category 2: Surgical management | (((((TS=(Surgical management )) OR TS=(Simple Prostatectomy)) OR TS=(Minimally invasive surgery )) OR TS=(Transurethral resection of the prostate )) OR TS=(TURP)) OR TS=(Transurethral Incision of the Prostate) |
| Category 3: Medical management | ((((((((((((((((((((((((((TS=(Medical management)) OR TS=(Alpha blocker)) OR TS=(α-blocker)) OR TS=(terazosin)) OR TS=(Hytrin)) OR TS=(doxazosin)) OR TS=(Cardura)) OR TS=(tamsulosin)) OR TS=(Flomax)) OR TS=(alfuzosin )) OR TS=(Uroxatral)) OR TS=(silodosin)) OR TS=(Rapaflo)) OR TS=(5-alpha-reductase inhibitors)) OR TS=(5-ARI)) OR TS=(Finasteride )) OR TS=(Proscar)) OR TS=(dutasteride)) OR TS=(Avodart)) OR TS=(Phosphodiesterase-5 inhibitor)) OR TS=(PDE inhibitor)) OR TS=(Tadalafil)) OR TS=(MTOPS)) OR TS=(medical therapy of prostatic symptoms)) OR TS=(Uroflow obstruction)) OR TS=(uroflow prostate)) OR TS=(therap* ) |
| Category 4: Advanced equipment | (((((((((((((TS=(Water vapor Thermal Therapy)) OR TS=(REZUM)) OR TS=(Laser Prostate)) OR TS=(Transurethral laser therapies)) OR TS=(Holmium laser enucleation of the prostate)) OR TS=(HoLEP)) OR TS=(thulium laser enucleation of the prostate )) OR TS=(Laser Photo-Vaporization of the Prostate )) OR TS=(Transurethral Vaporization of the Prostate )) OR TS=(Photoselective Vaporization of the Prostate)) OR TS=(Robotic Waterjet Treatment)) OR TS=(Prostatic artery embolization)) OR TS=(urolift)) OR TS=(Prostatic urethral lift ) |
| Category 5: Urologic terms | ((TS=(Urology)) OR TS=(Endourology)) OR TS=(Urologic) |

*TS, topic field tag searching title, abstract, and author keywords. Trailing spaces and capitalization are reproduced as entered.*

Query construction

Categories were combined with the Boolean operator AND to assemble four search queries. Every query included Category 1 and Category 5, so that every retrieved record contained both a BPH term and a urologic term. This was a deliberate design decision intended to restrict the corpus to literature published for and circulating within the urologic community, and its consequences are addressed in the Limitations.

Query 1, surgical management: Category 1 AND Category 2 AND Category 5.

Query 2, medical management: Category 1 AND Category 3 AND Category 5.

Query 3, advanced equipment: Category 1 AND Category 4 AND Category 5.

Query 4, combined surgical and advanced equipment: Category 1 AND Category 2 AND Category 4 AND Category 5.

<u>Database searches and record retrieval</u>

Web of Science was searched in February 2024. Altmetric Explorer was searched in February 2024 using PubMed-equivalent queries with the Title/Abstract field tag, because Altmetric Explorer does not accept Web of Science field tags. The 100 highest-ranked records per Web of Science query were exported, yielding 400 records and 208 unique records after duplicate removal. The four Altmetric Explorer queries returned 59, 30, 190 and 88 records respectively, yielding 367 records and 252 unique records after deduplication. The citation census date was 22 May 2024. Open-access status was determined by digital object identifier lookup in Unpaywall in August 2026 for the citation arm and from Altmetric Explorer for the attention arm.

<u>Screening</u>

Records were uploaded to Rayyan for duplicate removal and blinded screening. Three reviewers (A.R., K.M., N.Q.) independently screened titles and abstracts. An article was retained when at least two reviewers approved it, with discrepancies resolved by consensus and adjudication by the senior author. The citation arm was restricted to primary research articles in English. The attention arm applied deliberately broader publication-type criteria, permitting systematic reviews, meta-analyses, and narrative reviews. All other exclusions, including animal, in vitro, pediatric, and prostate malignancy studies, were applied identically to both arms.

<u>Derived variables</u>

The Adjusted Citation Index, reported in **Supplementary Table S1**, was defined as total citations divided by the number of years since publication, and accounts for differential exposure time across the citation arm. It was not used in any between-arm comparison

**Supplementary Table S1. Descriptive characteristics of the 50 most-cited BPH articles.**

| Characteristic | Value | Citations, median [IQR] |
|---|---|---|
| **Citation metrics** | | |
| Total citations, median [IQR] | 152 [106–211] | — |
| Total citations, range | 79–563 | — |
| Total citations, sum | 8,621 | — |
| Adjusted Citation Index, median [IQR] | 10.2 [6.2–14.4] | — |
| Year of publication, median [IQR] | 2008 [2003–2012] | — |
| Publication span | 1993–2018 | — |
| **Five-year period, n (%)** | | |
| 1991–1995 | 1 (2) | 329 |
| 1996–2000 | 7 (14) | 152 [114–179] |
| 2001–2005 | 6 (12) | 105 [86–201] |
| 2006–2010 | 19 (38) | 192 [96–244] |
| 2011–2015 | 15 (30) | 125 [111–160] |
| 2016–2020 | 2 (4) | 182 [169–196] |
| **Journal, n (%)** | | |
| European Urology | 29 (58) | 160 [104–224] |
| Urology | 16 (32) | 122 [105–178] |
| All other journals | 5 (10) | 169 [100–370] |
| Journal impact factor, median [IQR] | 6.5 [2.2–10.5] | — |
| **Study design, n (%)** | | |
| Randomized controlled trial | 29 (58) | 175 [118–231] |
| Retrospective/observational | 16 (32) | 124 [95–206] |
| Prospective | 5 (10) | 105 [82–111] |
| **Impact factor category, n (%)** | | |
| High impact (IF >10) | 16 (32) | 156 [122–212] |
| Medium impact (IF 5–10) | 13 (26) | 192 [100–238] |
| Moderate impact (IF 2–<5) | 19 (38) | 120 [94–179] |
| Low impact (IF <2) | 2 (4) | 185 [114–256] |
| **Country of first author, n (%)** | | |
| United States | 15 (30) | — |
| Italy | 5 (10) | — |
| Switzerland | 5 (10) | — |
| United Kingdom | 4 (8) | — |
| **Correlations (Spearman)** | | |
| Publication year × impact factor | $\rho = 0.738$ | $p < 0.001$ |
| Citations × impact factor | $\rho = 0.098$ | $p = 0.50$ |
| Citations × publication year | $\rho = -0.120$ | $p = 0.41$ |
| Adjusted Citation Index × impact factor | $\rho = 0.602$ | $p < 0.001$ |

*Per institutional preference, individual authors and institutions are not reported. Citation counts are Web of Science totals at the census date of 22 May 2024. The Adjusted Citation Index is defined in the Supplementary Methods. IF, impact factor; IQR, interquartile range.*

**Supplementary Table S2. Attention data for the 13 citation-arm articles retrievable in the Altmetric export.**

| Article | Year | Citations | Attention score | Open access | Digital Object Identifier |
|---|---|---|---|---|---|
| Comparison of prostatic artery embolisation (PAE) versus transurethral resection of the prostate (TURP) for benign prostatic hyperplasia: randomised, open label, non-inferiority trial | 2018 | 169 | 86 | Yes | 10.1136/bmj.k2338 |
| Management of acute urinary retention: a worldwide survey of 6074 men with benign prostatic hyperplasia | 2012 | 85 | 20 | Yes | 10.1111/j.1464-410x.2011.10430.x |
| Perioperative Outcomes of Robotic and Laparoscopic Simple Prostatectomy: A European-American Multi-institutional Analysis | 2015 | 123 | 15 | Yes | 10.1016/j.eururo.2014.11.044 |
| Tolterodine and tamsulosin for treatment of men with lower urinary tract symptoms and overactive bladder: a randomized controlled trial | 2006 | 410 | 12 | No | 10.1001/jama.296.19.2319 |
| Prostate-specific antigen as an estimator of prostate volume in the management of patients with symptomatic benign prostatic hyperplasia | 2003 | 79 | 11 | No | 10.1016/s0302-2838(03)00384-1 |
| Long-term effects of finasteride in patients with benign prostatic hyperplasia: a double-blind, placebo-controlled, multicenter study. PROWESS Study Group | 1998 | 152 | 10 | No | 10.1016/s0090-4295(98)00094-6 |
| A randomised trial comparing holmium laser enucleation versus transurethral resection in the treatment of prostates larger than 40 grams: results at 2 years | 2006 | 210 | 9 | No | 10.1016/j.eururo.2006.04.002 |
| The Hytrin Community Assessment Trial study: a one-year study of terazosin versus placebo in the treatment of men with symptomatic benign prostatic hyperplasia. HYCAT Investigator Group | 1996 | 175 | 6 | No | 10.1016/s0090-4295(99)80409-9 |
| Relationship of symptoms of prostatism to commonly used physiological and anatomical measures of the severity of benign prostatic hyperplasia | 1993 | 329 | 3 | No | 10.1016/s0022-5347(17)35482-4 |
| Holmium laser resection of the prostate: preliminary results of a new method for the treatment of benign prostatic hyperplasia | 1996 | 217 | 3 | No | 10.1016/s0090-4295(99)80381-1 |
| Open prostatectomy for benign prostatic enlargement in southern Europe in the late 1990s: a contemporary series of 1800 interventions | 2002 | 179 | 3 | No | 10.1016/s0090-4295(02)01860-5 |
| 180-W XPS GreenLight laser therapy for benign prostate hyperplasia: early safety, efficacy, and perioperative outcome after 201 procedures | 2012 | 122 | 1 | No | 10.1016/j.eururo.2011.11.041 |
| Dilutional hyponatremia of TURP syndrome: a historical event in the 21st century | 2004 | 88 | 0 | No | 10.1016/j.urology.2004.03.023 |

*Attention data were retrievable for 13 of the 50 citation-selected articles; the remaining 37 were not returned by the PubMed-converted Altmetric Explorer queries. Non-retrieval reflects query matching rather than absence of online attention, and no values were imputed for the remaining 37 articles. Articles are ordered by descending Altmetric Attention Score. DOI, digital object identifier.*

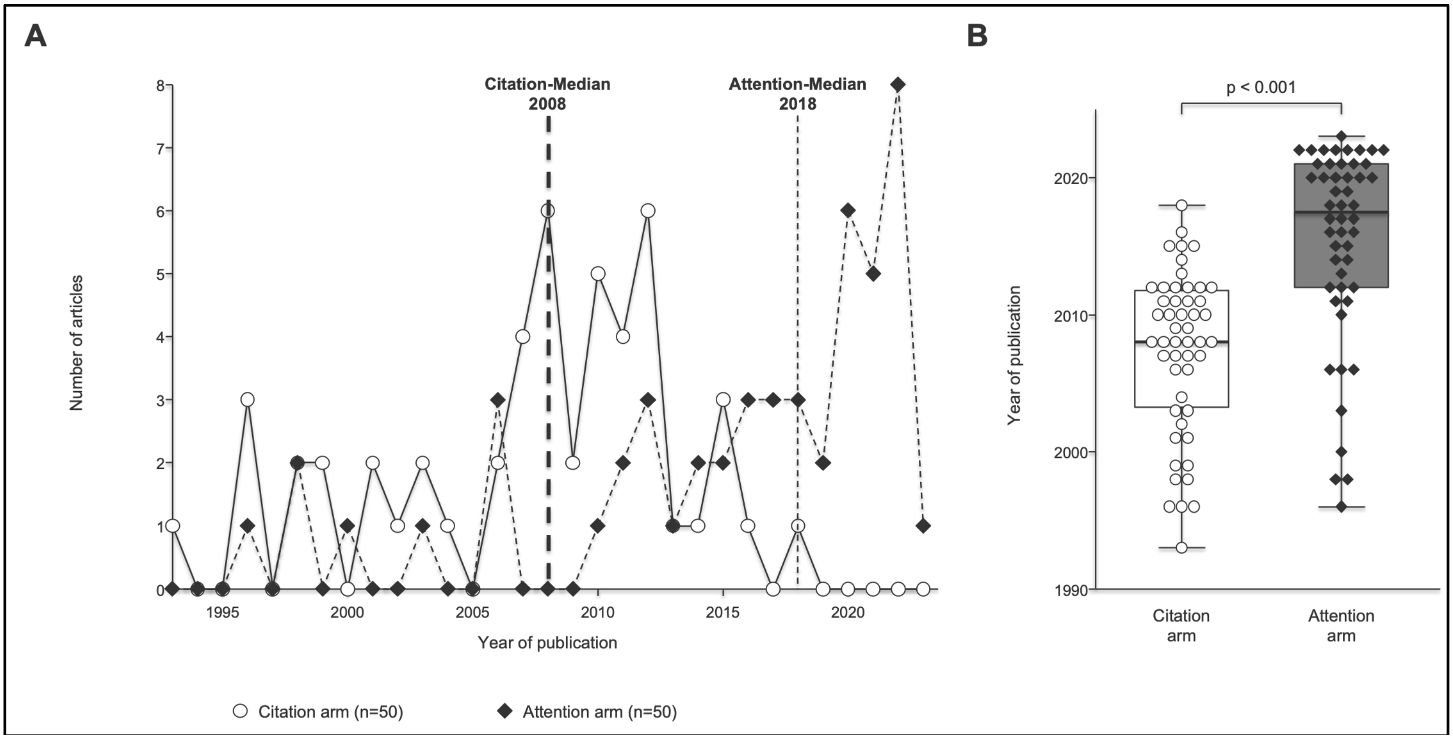


**Supplementary Figure S1. Publication year distribution of the two arms.**

(A) Number of articles by year of publication. Dotted lines mark the median year of each arm. (B) Distribution of publication year by arm, compared with the Mann-Whitney U test. Citation arm n = 50; attention arm n = 50.
